# Recent cannabis use and self-rated health in young and middle-aged adults: a propensity score-weighted analysis of the National Health and Nutrition Examination Survey (NHANES)

**DOI:** 10.64898/2026.08.20.26360898

**Authors:** Calvin Diep, Brittany N. Rosenbloom, Akash Goel, Rachael Bosma, Duminda N. Wijeysundera, Hance A. Clarke, Karim S. Ladha

**Affiliations:** Department of Anesthesiology and Pain Medicine, University of Toronto, Toronto, ON; Institute of Health Policy, Management and Evaluation, Dalla Lana School of Public Health, University of Toronto, Toronto, ON; Toronto Academic Pain Medicine Institute, Women’s College Hospital, Toronto, ON; Department of Anesthesia, Unity Health Toronto, Toronto, ON; Faculty of Dentistry, University of Toronto, Toronto, ON; Department of Anesthesia and Pain Management, University Health Network, Toronto, ON

**Author notes:** **Correspondence:** Calvin Diep, Department of Anesthesiology and Pain Medicine, 123 Edward St. (12^th^ floor), Toronto, ON, M5G 0A8.

**Keywords:** cannabis, self-rated health, quality of life, mental health

## Abstract

**Introduction:** Self-rated health is an important patient-centred measure of health. The relationship between cannabis use and self-rated health has been previously studied, although with methodologic concerns which we aimed to address in this investigation.

**Methods:** Propensity score weighted analyses of the National Health and Nutrition Examination Survey (NHANES) 2009–2018 were conducted. The primary exposure was self-reported cannabis use in the 30 days prior to survey response. The primary outcome was self-rated health measured on a five-level ordinal scale. Secondary outcomes included the number of days in the past months with: i) poor physical health, ii) poor mental health, and iii) activity limitations related to poor health. A weighted proportional odds regression model was used for the primary analysis and weighted zero-inflated negative binomial regression models were used for each secondary analysis.

**Results:** Among 22,055 adults aged 20–59 responding to the NHANES cannabis questionnaire, 14.4% endorsed use in the past 30 days. After reweighting the sample to balance cannabis users and non-users across sociodemographic, medical, and lifestyle characteristics, there was no statistically significant association between recent cannabis use and higher levels of self-rated health (OR 0.90, 95% CI 0.80–1.01). Cannabis use was associated with poor mental health and activity limitations in the past month, but not poor physical health.

**Conclusions:** Recent cannabis use was not associated with self-rated health but was associated with poor mental health and activity limitations in the past month. Cannabis users at risk of poor mental health should be connected with clinicians to help guide therapy.

## BACKGROUND

Cannabis use continues to increase in prevalence across the United States related to widespread decriminalization and social acceptance.^1^ This trend necessitates a comprehensive understand of the potential health impacts of cannabis use. Emerging evidence has demonstrated therapeutic efficacy for select indications such as cancer-related or non-cancer chronic pain, and possibly sleep issues.^2,3^ However, the literature to-date has mixed findings regarding the associations between cannabis use and holistic measures of health such as general health-related quality of life (HRQoL).^4,5^ Furthermore, few studies have examined the relationship between cannabis use and self-rated health, which has been shown to be an important patient-centred outcome and a valid proxy measure of HRQoL.^6^ As self-rated health is a strong predictor of morbidity, healthcare utilization, and mortality,^7–9^ elucidating accurate associations with cannabis use may provide insight into the broader public health implications of cannabis legalization and use.

The existing literature demonstrate mixed associations between cannabis use and self-rated health.^10,11^ There are also important limitations when interpreting these findings. For example, studies have generally classified participants as being cannabis users if they reported any use in the past year, which may be temporally distant (and thus causally unrelated) from the time of participation in the study.^11^ In addition, self-rated health has been operationalized as dichotomous, rather than ordinal or continuous response variables, leading to losses in important information.^10^ Therefore, the primary aim of this present study was to bridge these knowledge gaps by evaluating the association between cannabis use and self-rated health using more robust analytic methods. We hypothesized that recent cannabis use would be associated with worse self-rated health.

## METHODS

### Study setting and population

Data for this study were obtained from the National Health and Nutrition Examination Survey (NHANES), which is a cross-sectional survey designed by the National Center for Health Statistics (NCHS) and Centers for Disease Control and Prevention (CDC) and administered on a two-year cycle. The NHANES data collection protocols are approved by the NCHS Ethics Review Board and all survey participants provide informed consent prior to being interviewed and examined.

For this study, a data set was constructed using publicly available files from seven two-year cycles of NHANES responses (2005–06, 2007–08, 2009–10, 2011–12, 2013–14, 2015–16, and 2017–18). These cycles were selected as they are contiguous and contain questionnaire data related to cannabis use needed to define our exposure. The study population consisted of all respondents to the NHANES cannabis questionnaire, which was offered to adults 20 to 59 years of age.

### Outcomes

The outcomes in this study were the four items of the CDC’s HRQoL-4 questionnaire, also known as the Healthy Days core module. The primary outcome was self-rated general health status, with five possible responses on a five-level ordinal scale: “poor”, “fair”, “good”, “very good”, or “excellent”. This single item has demonstrated validity as a proxy measure of HRQoL and is simple to administer, easy for subjects to respond to, and captures both the physical and mental domains of health.^6^ Data for this item was available in seven survey cycles from 2005–06 to 2017–18. Secondary outcomes were: i) days with poor physical health (defined as physical illness or injury) in the past 30 days, ii) days with poor mental health (defined as stress, depression, or problems with emotions) in the past 30 days, and iii) days with activity limitation due to poor health (physical or mental) in the past 30 days. Data for secondary outcomes was only available in the four cycles from 2005–06 to 2011–12.

### Exposure

The primary exposure was recent cannabis use. Survey respondents were classified as recent cannabis users if they reported using cannabis at least once in the 30 days prior to survey administration. This is consistent with commonly used definitions in population-based surveys.^12,13^ The comparator group were those with no reported cannabis use in the past 30 days.

### Covariates

Demographic characteristics included age, biological sex, and race/ethnicity (white, Black, or other), marital status (never married, currently married, widowed/divorced/separated), and number of other people living in same household (none, 1–4, ≥5). Socioeconomic factors included education (secondary school or beyond), employment status, number of hours worked per week (<20, 21–40, 41–80, >80), family income-to-poverty line ratio (<1, 1–3, or ≥3), level of food security (full, marginal, low), health insurance coverage, and access to routine primary care. Medical characteristics included hypertension, diabetes, HbA1c (%), body mass index (BMI, in kg/m^2^), coronary artery disease, heart failure, stroke or TIA, anemia, hemoglobin concentration (dL/g), renal disease, asthma, chronic obstructive pulmonary disease, liver disease, thyroid disease, arthritis, history of cancer, and any hospitalization in the past year. Neuropsychiatric characteristics included current depressive symptoms (measured using the Patient Health Questionnaire [PHQ-9]; ordinal responses), confusion or memory problems, and having seen a mental health professional in the past year. Medication-related characteristics were polypharmacy (defined as ≥4 prescription medications in the past month), opioid prescriptions in the past month, and other pain medication prescriptions in the past month. Health-related lifestyle factors included number of hours of sleep per night (adequate 6–9, insufficient <6, or excess >9), diagnosed sleep disorder, self-reported poor diet, number of non-homemade meals per week, number of days per week with ≥10 minutes of moderate-to-vigorous physical activity, number of sedentary minutes per day, smoking status, any smoking inside home, heavy drinker (classified as ≥2 drinks per night on average). Survey cycle (2005–06, 2007–08, 2009–10, 2011–12, 2013–14, 2015–16, and 2017–18) was also considered as a covariate.

### Statistical analyses

Baseline characteristics of the exposure groups were summarized using means with standard deviations (or medians with interquartile ranges, as appropriate) or counts with proportions. Propensity scores were generated using multivariable logistic regression modeling including all listed covariates. Two separate propensity score models were developed: one with data from all seven survey cycles (2005 to 2018) for modeling the primary outcome, and another with data from 2005 to 2012 for modeling secondary outcomes. Stabilized inverse probability of treatment (IPT) weights targeting the average marginal treatment effect (ATE) were derived from propensity scores and incorporated into outcome models. Therefore, for all outcomes, crude (i.e., unweighted and unadjusted) models were fitted, followed by weighted models using IPT weights and robust sandwich-type variance estimators to account for within-subject correlation induced by weighting.

For the primary outcome of self-rated health, an ordinal (i.e., proportional odds) logistic regression model was used. Model estimates were presented as odds ratios (ORs) with 95% confidence intervals, interpreted as the relative odds of a *higher* (i.e., better) level of self-rated health compared to reference groups. For each of the secondary outcomes, a weighted zero-inflated negative binomial (ZINB) model was fitted, given the expectation for zero-inflation and overdispersion of values not fitting a Poisson distribution. ZINB models have two parts: 1) a logistic regression model evaluating the log odds of a response value of zero, and 2) a negative binomial regression model evaluating the incident rate of the count amongst those with non-zero values. Model estimates from the first part of the ZINB model were reported as ORs with 95% confidence intervals and interpreted as the relative *odds of reporting zero days* compared to reference groups. Model estimates from the second part of the ZINB model were reported as incident rate ratios (IRRs) with 95% confidence intervals and interpreted as the relative *increase in the incident rate of number of days* in the past month, compared to reference groups.

Several subgroup analyses were conducted, stratifying the cohort by the following variables: i) age (dichotomized into young adults aged 20–39 years or middle-aged adults aged 40–59 years), ii) prescription of opioid-containing medication in the past 30 days and iii) prescription of non-opioid pain medications in the past 30 days. For each subgroup analysis, the propensity model was re-specified to generate IPT weights which balanced baseline characteristics between cannabis users and non-users before incorporation in outcome models.

Observations with missing exposure or outcome data were excluded. For covariates with ≥5% missingness, an additional indicator variable was added to retain these observations in analyses. Statistical significance was defined as a two-sided p-value <0.05. Analyses were performed with R version 4.2 (R Core Team, Vienna, Austria).

## RESULTS

The NHANES cannabis questionnaire was administered to 26,269 adults across the United States aged 20–59 years between 2005 and 2018. After excluding those with missing cannabis data (4214; 16.0%), 22,055 respondents were retained. There were 3177 (14.4%) respondents who endorsed cannabis use in the past 30 days, and 18,878 (85.6%) who did not. Recent cannabis users and non-users were different across many baseline demographic, socioeconomic, medical, and lifestyle characteristics (Table 1). Standardized mean differences between the cohorts were reduced to ≤10% across all covariates after the sample was weighted by IPT.

**Table 1.**
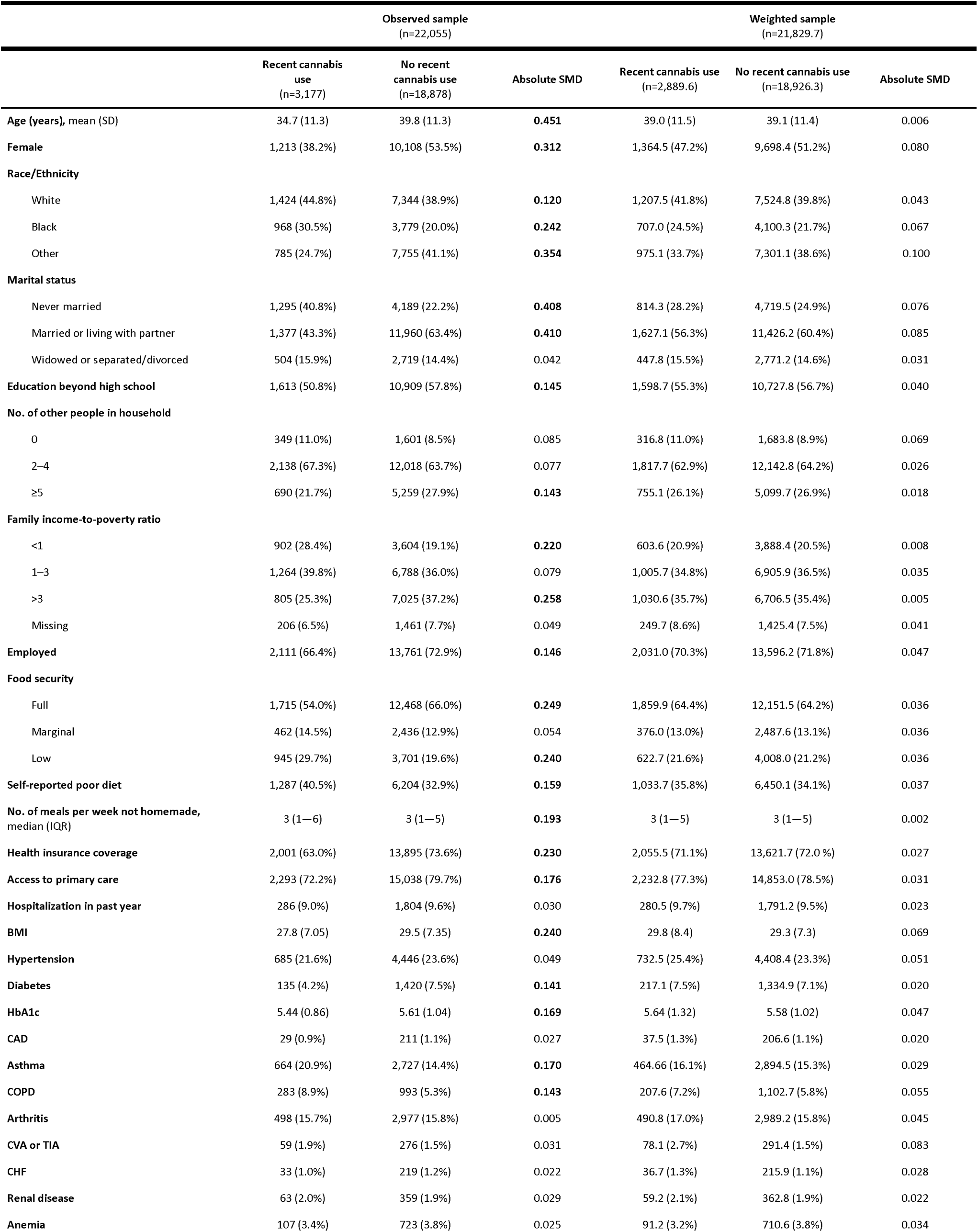
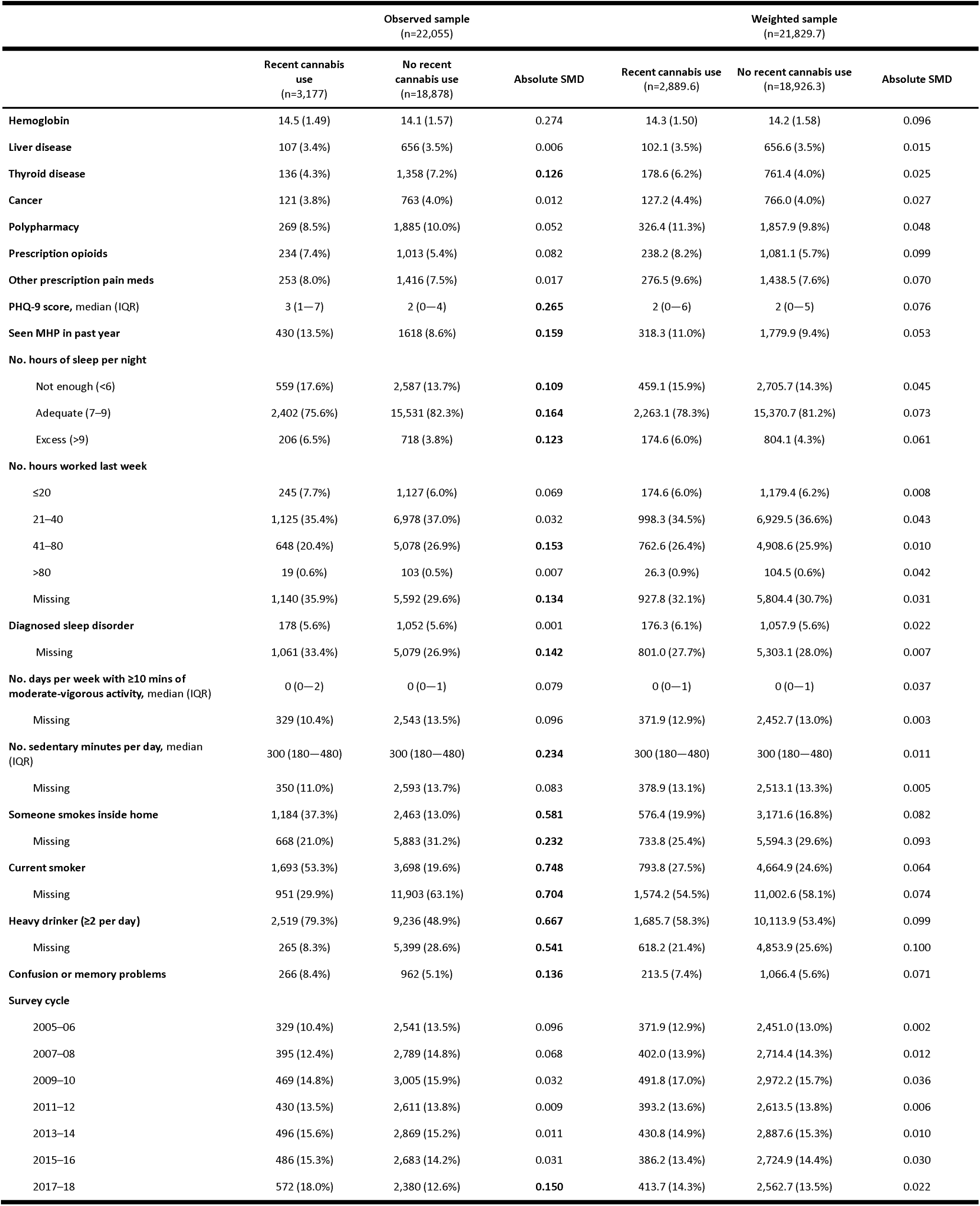
Cohort characteristics before and after weighting by inverse probability of exposure.

The distribution of self-rated health levels in the observed (unweighted) sample are displayed in Table 2. In the crude (unweighted and unadjusted) proportional odds logistic regression model, cannabis use was associated with 15% lower odds of better self-rated health (OR 0.85, 95% CI 0.79–0.91). This association was no longer significant in the weighted sample (OR 0.90, 95% CI 0.80–1.01). Secondary outcomes between cannabis users and non-users are displayed in Table 2. In the weighted ZINB models for secondary outcomes, cannabis use was associated with lower odds of having zero days with poor mental health in the past month (OR 0.63, 95% CI 0.48–0.83) and zero days limited by poor health (OR 0.62, 95% CI 0.47–0.82), but not for poor physical health (Table 3). In the negative binomial portion of the models, cannabis use was not associated with the incident rate of greater number of days of poor physical health, mental health, or days with activities limited by poor health.

**Table 2.** Outcomes for recent cannabis users in the observed (unweighted) sample.

|  | **Recent cannabis use** | **No recent cannabis use** |
| --- | --- | --- |
| <u>2005–2018</u> | n=3,177 | n=18,878 |
| **Self-rated health** |  |  |
| Poor | 122 (3.8%) | 614 (3.3%) |
| Fair | 573 (8.6%) | 3,174 (16.8%) |
| Good | 1220 (38.4%) | 6,864 (36.4%) |
| Very good | 893 (28.1%) | 5,338 (28.3%) |
| Excellent | 366 (11.5%) | 2,873 (15.2%) |
|  | **Recent cannabis use** | **No recent cannabis use** |
| <u>2005–2012</u> | n=1,623 | n=10,946 |
| **No. days physical health not good in past month** |  |  |
| Median (IQR) | 0 (0–4) | 0 (0–3) |
| Zeros (%) | 979 (60.3%) | 7,126 (65.1%) |
| Median (IQR) of non-zeros | 6 (3–15) | 5 (2–15) |
| **No. days mental health not good in past month** |  |  |
| Median (IQR) | 2 (0–10) | 0 (0–4) |
| Zeros (%) | 668 (41.2%) | 6,364 (58.1%) |
| Median (IQR) of non-zeros | 7 (3–15) | 5 (2–15) |
| **No. days activity limited by poor health in past month** |  |  |
| Median (IQR) | 0 (0–1) | 0 (0–0) |
| Zeros (%) | 1,166 (74.8%) | 8,825 (80.6%) |
| Median (IQR) of non-zeros | 5 (2–14) | 5 (2–14) |

**Table 3.** Associations between recent cannabis use and study outcomes.

|  | Higher levels<br>of self-rated<br>health<br>(2005–18) | Days of poor physical health<br>(2005–12) |  | Days of poor mental health<br>(2005–12) |  | Days limited by health<br>(2005–12) |  |
| --- | --- | --- | --- | --- | --- | --- | --- |
|  | PO model<br>OR (95% CI) | Zero model<br>OR (95% CI) | Count model<br>IRR (95% CI) | Zero model<br>OR (95% CI) | Count model<br>IRR (95% CI) | Zero model<br>OR (95% CI) | Count model<br>IRR (95% CI) |
| Crude | **0.85**<br>(0.79–0.91) | **0.80**<br>(0.70–0.91) | 1.07<br>(0.96–1.18) | **0.44**<br>(0.38–0.51) | **1.13**<br>(1.04–1.23) | **0.57**<br>(0.49–0.66) | 0.97<br>(0.84–1.12) |
| Weighted | 0.90<br>(0.80–1.01) | 0.93<br>(0.74–1.17) | 1.11<br>(0.90–1.37) | **0.63**<br>(0.48–0.83) | 0.96<br>(0.85–1.08) | **0.62**<br>(0.47–0.82) | 0.98<br>(0.70–1.38) |
| Subgroups: |  |  |  |  |  |  |  |
| Age |  |  |  |  |  |  |  |
| 20–39 | **0.81**<br>(0.70 – 0.93) | 1.04<br>(0.73 – 1.47) | 1.32<br>(0.94 – 1.87) | **0.59**<br>(0.39 – 0.89) | 0.99<br>(0.79 – 1.22) | 0.40<br>(0.02 – 6.47) | 0.63<br>(0.25 – 1.58) |
| 40–59 | 0.93<br>(0.76 – 1.13) | 0.84<br>(0.60 – 1.17) | 0.99<br>(0.78 – 1.26) | **0.61**<br>(0.41 – 0.89) | 0.95<br>(0.78 – 1.16) | **0.62**<br>(0.43 – 0.88) | 1.02<br>(0.74 – 1.41) |
| Rx opioids |  |  |  |  |  |  |  |
| Yes | 0.98<br>(0.42 – 2.30) | 1.05<br>(0.55 – 2.01) | 1.02<br>(0.83 – 1.25) | 0.63<br>(0.31 – 1.26) | 1.14<br>(0.84 – 1.55) | 0.85<br>(0.45 – 1.59) | 0.89<br>(0.69 – 1.15) |
| No | **0.87**<br>(0.77 – 0.99) | 0.90<br>(0.69 – 1.17) | 1.20<br>(0.89 – 1.63) | **0.59**<br>(0.43 – 0.82) | 0.94<br>(0.81 – 1.08) | **0.59**<br>(0.43 – 0.81) | 1.11<br>(0.67 – 1.84) |
| Other Rx pain<br>meds |  |  |  |  |  |  |  |
| Yes | 0.96<br>(0.66 – 1.41) | 0.93<br>(0.51 – 1.73) | 1.03<br>(0.82 – 1.29) | **0.45**<br>(0.22 – 0.93) | 1.10<br>(0.84 – 1.44) | 0.76<br>(0.43 – 1.36) | 0.91<br>(0.67 – 1.24) |
| No | **0.87**<br>(0.76 – 1.00) | 0.90<br>(0.70 – 1.17) | 1.19<br>(0.90 – 1.57) | **0.59**<br>(0.43 – 0.81) | 0.94<br>(0.81 – 1.09) | **0.57**<br>(0.40 – 0.80) | 1.03<br>(0.63 – 1.66) |

All estimates represent the relative association of the outcome for subjects reported recent cannabis use (i.e., in the past 30 days) compared to subjects with no recent cannabis use.

Results of the weighted subgroup analyses are reported in Table 3. Among young adults cannabis use was associated with lower odds of better self-rated health (OR 0.81, 95% CI 0.79– 0.93), whereas this association was not observed for middle-aged adults. Among those with recent prescriptions for opioid or non-opioid pain medications, there was no association between cannabis use and self-rated health. Conversely, for those without prescription pain medications, recent cannabis use was associated with worse self-rated health. For nearly all subgroups, cannabis use was associated with having at least one day of poor mental health in the past month (Table 3).

## DISCUSSION

In this propensity score-weighted analysis of NHANES respondents aged 20–59 from 2005 to 2018, recent cannabis use was not associated with level of self-rated health. However, there was a negative association for some subgroups including younger adults and those with no recent prescriptions for pain medications. Across all subgroups, recent cannabis use was associated with experiencing poor mental health in the past month.

Our primary findings conflict with those of Tsai *et al*. who had also evaluated the association between cannabis use and self-rated health using NHANES data, but with different definitions of the exposure and outcome.^10^ In their study, cannabis users were classified by whether they reported having ever used cannabis at least once per month for an entire year. However, for some subjects, the period of regular cannabis use may have been temporally distant from the time of reporting self-rated health. For example, for a middle-aged adult who last regularly used cannabis in their teenage years, the association between cannabis use and self-rated health would be confounded by many more factors than for another adult who more recently used cannabis. Furthermore, the self-rated health outcome variable was dichotomized as suboptimal (“poor” or “fair”) versus good, resulting in a substantial loss of information from the original five-level ordinal response options. While the adjusted odds of suboptimal health were 1.34 times greater (95% CI 1.05–1.69) amongst those with versus without a history of regular cannabis use in their study, this estimate is subject to indirectness and residual confounding bias. In addition to restricting our definition of cannabis users to those who had used in the past 30 days (i.e., more recent to the outcome measure), our study more accurately estimated the association between cannabis and self-rated health using propensity score methods to further reduce confounding and a more efficient analytic model that retained self-rated health as an ordinal – rather than dichotomous – response variable.^14,15^ Furthermore, our findings align with observational studies and randomized controlled trials which have reported mixed and inconclusive associations between cannabis and various measures of HRQoL.^4,5^

Recent cannabis use was associated with experiencing poor mental health in the past month in our study. This is consistent with previous studies showing negative associations between cannabis and mental domains of HRQoL measures.^16,17^ Cannabis is known to promote the onset or exacerbation of symptoms of anxiety, depression, or psychosis in some users, especially with repeated or long-term exposure.^18,19^ On the other hand, poor mental health has also been shown to promote incident cannabis use. That is, patients with untreated or refractory symptoms of depression, anxiety, or posttraumatic stress disorder often self-medicate using cannabis products as experimental therapy,^20^ despite the paucity of strong evidence or clinical guidelines to support this.^19,21^ Given the cross-sectional nature of this study data, the temporal (and causal) relationship between cannabis use and new mental health concerns could not be elucidated here, but have recently been reviewed elsewhere.^22–24^

As the evidence base continues to grow in support of the therapeutic benefits of cannabis for indications such as pain and sleep-related issues, greater attention should be paid to how these effects translate to outcomes that matter most to patients, such as changes in HRQoL and function. The relationships between cannabis and these patient-centred outcomes are nuanced and confounded by many factors not always captured in clinical studies. Investigations that capture granular data related to indications for use, products and formulations, and dose/frequency of use are needed so we can disentangle whether these factors contribute to poor health outcomes. Such real world evidence of cannabis users with repeated measures of pain, sleep, and mental health are valuable to clinicians when counseling patients about risks related to specific products or consumption behaviours.^25,26^

There are several limitations to consider in this study. First, data collected by the NHANES is cross-sectional and not appropriate for inferring causation. For instance, our findings do not imply that cannabis use causes poor mental health. Rather, there are likely differences between cannabis users and non-users across characteristics that are associated with mental health as well as behaviours that result from poor mental health. Second, our findings may be subject to residual confounding. Despite accounting for an extensive set of covariates in analyses, there are still others that may be relevant confounders such as medical and psychiatric factors that influence cannabis use and contribute to self-rated health. Third, data collected from surveys such as NHANES are voluntary and self-reported and thus subject to the “healthy volunteer” selection bias. Thus, only persons willing and able to participate in NHANES’ extensive questionnaires and examinations are included. Furthermore, the stigma associated with cannabis use is likely to have negatively impacted participants’ response rates and underestimated the true prevalence of use. Non-responders to the cannabis questionnaire are thus likely to be different from responders on various sociodemographic and medical characteristics. Future prospective studies reporting rates of non-response to cannabis questionnaires can incorporate quantitative bias analytic techniques to improve estimation of associations between cannabis and health-related outcomes.

## Conclusion

Recent cannabis use was not associated with better or worse levels of self-rated health in a sample of young and middle-aged adults across the United Status. Cannabis use was associated with experiencing poor mental health and activity limitations related to health in the past month, but not poor physical health in the past month. Real world data about pain, sleep, and mental health measures from cannabis users in future investigations can provide new insights on how these factors mediate changes in health-related quality of life.

## Acknowledgements

N/A

## Author contributions

- Calvin Diep: Conceptualization, Methodology, Software, Formal analysis, Investigation, Data Curation, Writing – Original Draft, Writing – Review & Editing
- Brittany Rosenblood: Methodology, Writing – Review & Editing
- Akash Goel: Methodology, Writing – Review & Editing
- Rachael Bosma: Methodology, Writing – Review & Editing
- Duminda Wijeysundera: Conceptualization, Methodology, Writing – Review & Editing
- Hance Clarke: Writing – Original Draft, Writing – Review & Editing, Supervision
- Karim Ladha: Conceptualization, Methodology, Writing – Original Draft, Writing – Review & Editing, Supervision

## Informed consent / patient consent

The NHANES data collection protocols are approved by the NCHS Ethics Review Board and all survey participants provide informed consent prior to being interviewed and examined. Therefore, additional research ethics approval was not sought from the local Institutional Review Board at Unity Health Toronto or University of Toronto.

## Data availability statement

NHANES data are publicly available from https://wwwn.cdc.gov/nchs/nhanes/

